# Mapping the Landscape of Anaemia Research in Pregnant and Postpartum Women: A Bibliometric Analysis & Topic Modelling Study

**DOI:** 10.1101/2025.10.04.25337319

**Authors:** Olalekan A. Uthman, Tabassum Firoz, María Barreix, Lisa M Rogers

## Abstract

**Background:** Maternal anaemia affects 35.5% of pregnant women globally and represents a persistent public health challenge with significant maternal and neonatal health consequences. Despite substantial research investment, progress towards anaemia reduction remains limited, suggesting potential misalignment between research priorities and evidence-based intervention needs. Understanding the knowledge structure and thematic evolution of maternal anaemia research is essential for identifying gaps and informing future research strategies and guidelines development.

**Objectives:** This study aimed to map the knowledge structure and evolution of research on anaemia in pregnant and postpartum women from 2000-2024, analyse global collaboration networks and geographic distribution, and identify emerging themes and research gaps using bibliometric analysis and topic modelling.

**Methods:** A comprehensive bibliometric analysis was conducted using Web of Science Core Collection database, encompassing 1,007 documents published between 2000-2024. Standard bibliometric indicators were calculated using VOSviewer and CiteSpace software. Latent Dirichlet Allocation topic modelling identified thematic clusters. Linear regression analysis examined temporal trends and geographic patterns.

**Results:** Publication output demonstrated exponential growth (R² = 0.9054), increasing from 12 documents in 2000 to 118 in 2022. Research concentrated in high-income countries (68.5% of publications) despite inverse disease burden distribution. Geographic disparities revealed substantial regional research gaps, particularly in Sub-Saharan Africa and South Asia. Topic modelling revealed nine clusters dominated by health outcomes research (77.4%), with less attention to prevention (20.8%) and diagnosis (6.3%).

**Conclusions:** Maternal anaemia research exhibits fundamental structural imbalances with geographic concentration contradicting disease burden patterns and thematic evolution away from prevention towards treatment focus. The virtual absence of implementation research represents a critical barrier to knowledge translation. Strategic research rebalancing prioritising implementation science, prevention research, and capacity building in high-burden regions is essential for achieving population-level anaemia reduction goals.

## Background

Maternal anaemia remains one of the most persistent global public health challenges affecting approximately 35.5% of pregnant women worldwide. The prevalence is as high as 52% in low- and middle-income countries, particularly in Sub-Saharan Africa and South Asia^1^. Despite decades of intervention efforts, global progress towards anaemia reduction has remained stagnant, with prevalence declining by only 1.6 percentage points between 2000 and 2016^3^, reflecting the complex interactions between nutritional deficiencies, obstetric causes, chronic and infectious diseases, and the broader socio-economic context.^2^

The health consequences of maternal anaemia include maternal and neonatal morbidity and mortality as well as intergenerational transmission of poor health outcomes^6^. Both direct and indirect causes of maternal deaths can be due to anaemia. The most recent systematic analysis of the global causes of maternal death shows that hemorrhage, for which anaemia cna be a risk factor, remains the leading cause of maternal deaths globally. The analysis also shows that there has been no substantial change in the proportion of maternal deaths from indirect causes since the two previous WHO analyses in 2006 and 2014.

The thematic focus of existing research on anaemia demonstrates fragmentation and misalignment with public health priorities. Research attention has mainly focused on nutritional supplementation strategies, particularly iron and folic acid interventions, whilst implementation science and health systems research remain underrepresented^7^. Within the maternal health continuum, the postpartum period has received disproportionately limited research attention despite evidence showing that anaemia can persist or even worsen during this time ^8^. Global estimates also show that most maternal deaths occur in the postpartum period signifying the need for more attention during this critical period.

In May 2023, WHO launched a [Comprehensive framework for action to accelerate anaemia reduction](https://www.who.int/publications/i/item/9789240074033), advocating for coordinated action across systems and emphasizing a broad approach to diagnosis, prevention and management. This includes addressing all main causes of anaemia and the broader social inequities related to education, poverty, food insecurity and lack of access to family planning, health and nutrition services and clean water, sanitation and hygiene. Addressing both the causes and risk factors simultaneously is essential for effective anaemia control.

The complexity and fragmentation of the maternal anaemia research landscape necessitate comprehensive evidence mapping approaches to characterise knowledge structures, identify research gaps, and inform research priorities for guideline development. Bibliometric analysis provides powerful methodological approaches for synthesising large research corpora, enabling identification of collaboration patterns, thematic evolution, and geographic distribution of research activity^9^. Topic modeling techniques, particularly Latent Dirichlet Allocation, offer sophisticated approaches for identifying latent thematic structures within extensive literature collections, facilitating objective assessment of research priorities and trends^10^.

Previous bibliometric analyses in related maternal health domains have demonstrated the utility of these approaches for identifying research inequities, collaboration patterns, and emerging research frontiers^12^. However, no comprehensive bibliometric analysis has specifically examined the landscape of anaemia research in pregnant and postpartum women using contemporary computational methods integrated with policy-relevant frameworks.

The aim of this study is to address these knowledge gaps through comprehensive bibliometric analysis and topic modelling of anaemia research in pregnant and postpartum women by: (1) mapping the knowledge structure and evolution of research on anaemia in pregnant and postpartum women over the past 25 years; (2) analysing geographic distribution and global collaboration networks of research activity to identify patterns of research concentration and underrepresentation; and (3) uncovering emerging research themes and declining areas through temporal trend analysis and topic modelling.

## Methods

### Study Design

This research employed a comprehensive bibliometric analysis and topic modelling approach to systematically map the intellectual landscape of anaemia research in pregnant and postpartum women from 2000 to 2024 (**Figure 1**). The study integrated quantitative bibliometric indicators with qualitative content analysis through advanced computational techniques, including network analysis, topic modelling, and temporal trend assessment. This mixed-methods design enabled examination of research productivity patterns, collaboration networks, thematic evolution, and geographic distribution. Figure 1 provides an overview of the methodology. Each step is discussed in detail in Appendix 1.

**Figure 1:**
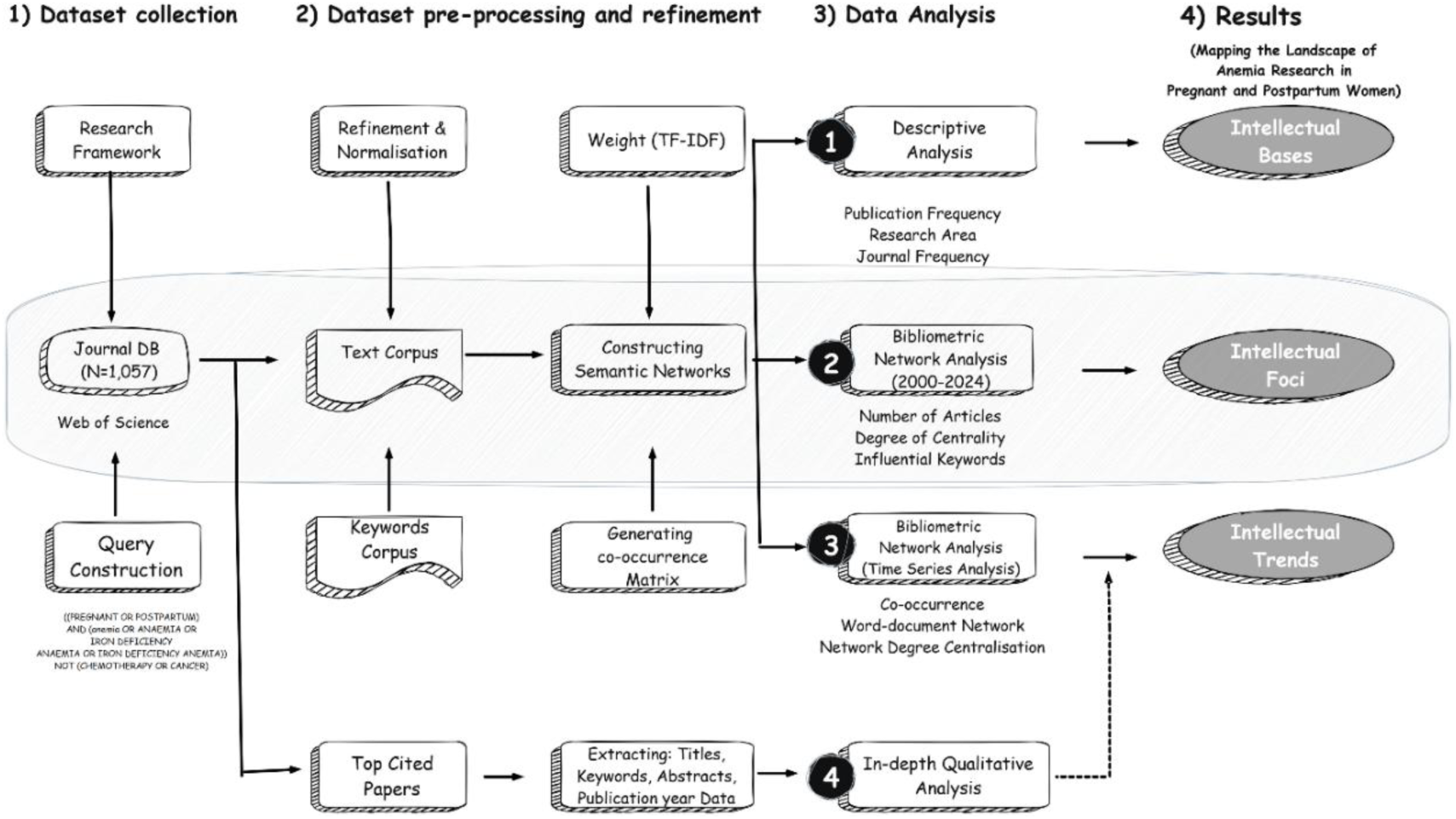
Overview of the methodology.

We additionally performed manual classification of all included studies against the categories for guideline development to serve as a reference standard (‘ground truth’) for evaluating the distribution of machine-generated topics. Two reviewers independently assigned categories with disagreements resolved by consensus. An additional 50 studies were excluded at this stage.

### Interactive Dashboard Development

A comprehensive interactive dashboard was developed and deployed to facilitate detailed exploration of the bibliometric analysis findings (https://maternalanemia.streamlit.app/) (**A ppendix 1**). This web-based platform provides stakeholders with dynamic access to the complete analytical dataset through multiple interconnected visualisation modules designed for different user needs and expertise levels. The dashboard comprises several key analytical sections enabling comprehensive data exploration. Advanced analytical tools include an Authors Discovery module for identifying influential researchers and collaboration patterns, and a Topics Discovery section providing detailed thematic analysis with interactive topic model exploration.

## Results

The bibliometric analysis encompassed 1,007 documents on anaemia in pregnant and postpartum women published between 2000 and 2024 (**Figure 2**). The dataset comprised 814 research articles (80.8%), 110 reviews (10.9%), 51 meeting abstracts (5.1%), 18 proceedings papers (1.8%), and smaller proportions of other document types including editorials, corrections, and news items.

**Figure 2:**
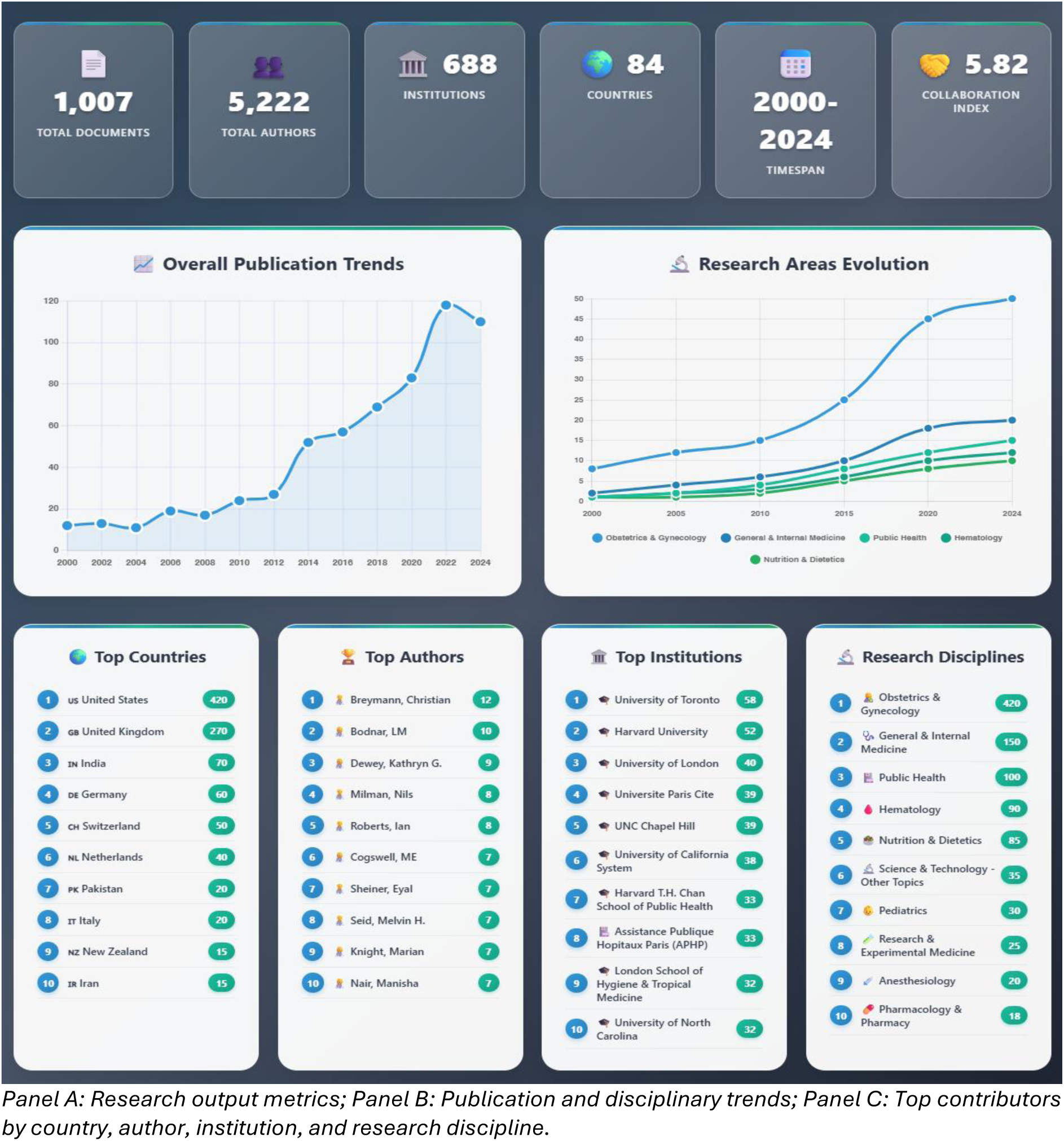
Overview of bibliometric analysis of anaemia research in pregnant and postpartum women (2000-2024). Panel A: Research output metrics; Panel B: Publication and disciplinary trends; Panel C: Top contributors by country, author, institution, and research discipline.

### Publication Trends

Publications originated from 84 countries across 688 institutions and were disseminated through 363 distinct sources.

### Temporal Publication Patterns

Annual publication output exhibited a marked exponential growth trajectory over the 25-year study period, with an R² value of 0.9054 indicating strong model fit (**Figure 2**). Publications increased from 12 in 2000 to a peak of 118 in 2022, representing a nearly ten-fold increase. The trend demonstrated particular acceleration from 2013 onwards, with annual output rising from 35 publications in 2013 to sustained levels above 50 publications from 2015. Average annual publication output across the entire timespan was 40.28 documents per year. The growth pattern reflects increased scholarly attention to maternal anaemia in pregnancy research in recent years.

### Terminology Landscape and Keyword Evolution

The most frequently occurring multi-word phrases demonstrated clear thematic clustering around key research domains (**Figure 3**). Maternal-related terms dominated the largest segment, encompassing phrases such as “maternal mortality,” “maternal health,” and “maternal outcomes.” The postpartum domain represented another substantial cluster, including terms like “postpartum haemorrhage” and “postpartum depression.” Pregnancy-related terminology was represented through phrases including “preterm birth,” “caesarean section,” and gestational periods (“third trimester”). Haematological parameters constituted another cluster, featuring “haemoglobin levels,” “blood loss,” and “blood transfusion.” The frequency distribution, indicated by colour intensity, showed maternal-related phrases achieving the highest occurrence rates, followed by clinical intervention and diagnostic terminology (Figure 3).

**Figure 3:**
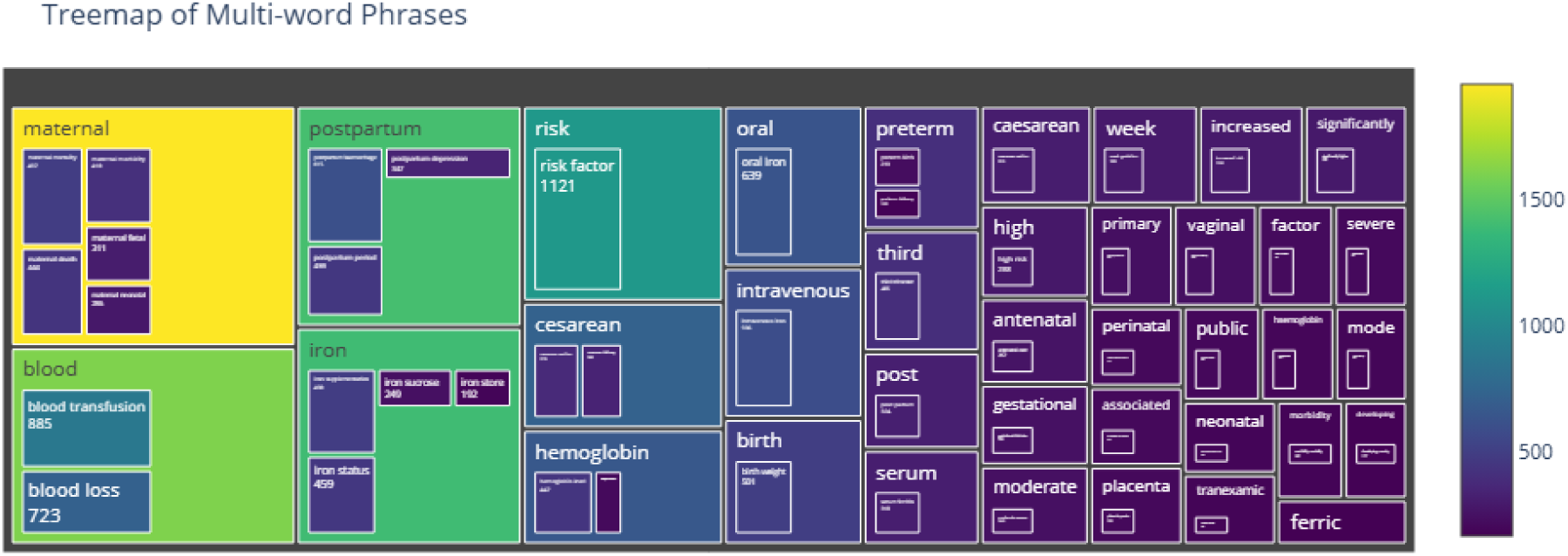
Terminology landscape.

The keyword evolution analysis spanning 2000-2024 demonstrated dynamic shifts in research terminology and focus areas (**Figure 4**). Early years (2000-2003) were characterised by fundamental terminology including “anaemia,” “pregnancy,” and basic haematological markers. The period from 2004-2007 showed expansion into “iron deficiency” and “folate” supplementation research. A notable transition occurred from 2008-2010, with the emergence of “iron supplementation” as a prominent keyword, alongside increased attention to “outcomes” and “prevalence” studies. The period 2011-2013 marked the introduction of more sophisticated clinical terminology, including specific treatment modalities and “India” as a geographical focus, reflecting increased research activity in high-burden regions.

**Figure 4:**
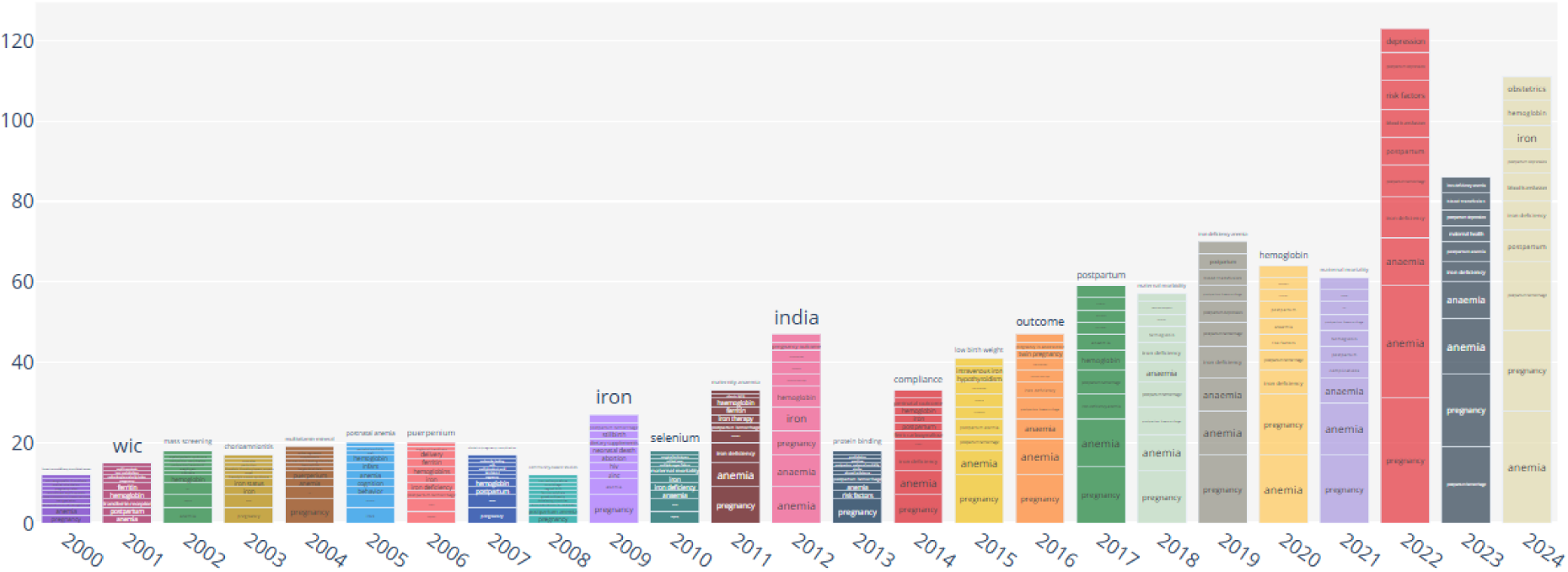
Evolution of keywords.

The years 2014-2016 demonstrated diversification into “pregnancy outcomes,” “anaemia prevalence,” and “prevention” strategies. A significant evolution occurred in 2017-2019, with the emergence of “depression,” “gestational diabetes,” and “quality of life” keywords, indicating research expansion beyond traditional haematological parameters into broader maternal health and wellbeing outcomes.

The most recent period (2020-2024) revealed further specialisation with keywords including “systematic review,” “meta-analysis,” “randomised controlled trial,” and “machine learning,” suggesting methodological advancement and evidence synthesis approaches. Geographic diversification continued with keywords representing various regions and populations, whilst clinical terminology evolved to include specific biomarkers and advanced therapeutic approaches.

### Research Discipline Distribution and Trends

Research discipline analysis revealed the multidisciplinary nature of maternal anaemia research with clear domain predominance. Obstetrics & Gynaecology represented the largest disciplinary contribution with 420 publications (41.7%), confirming the clinical focus on maternal health. General & Internal Medicine contributed 150 publications (14.9%), whilst Public Health accounted for 100 publications (9.9%). Haematology generated 90 publications (8.9%), and Nutrition & Dietetics contributed 85 publications (8.4%). Other disciplines included Science & (3.5%), Paediatrics (3.0%). Research & Experimental Medicine (2.5%), Anaesthesiology (2.0%), and Pharmacology & Pharmacy (1.8%). The research discipline trends over time demonstrated dynamic shifts in disciplinary engagement from 2000-2024. Obstetrics & Gynaecology showed consistent growth throughout the period, with notable acceleration from 2015 onwards, reaching peak output exceeding 50 publications annually by 2022. General & Internal Medicine displayed moderate but steady growth, whilst Public Health contributions increased markedly after 2010.

### Geographic Distribution and Country Networks

Country-level analysis revealed significant geographic concentration in maternal anaemia research productivity (**eFigure 3**). The United States dominated with 420 publications (41.7%), followed by the United Kingdom with 270 publications (26.8%). India contributed 70 publications (7.0%), representing the leading contribution from low- and middle-income countries.

The country collaboration network visualisation displayed extensive global research partnerships with notable regional clustering patterns. The network showed the United States and United Kingdom as central nodes with multiple international connections. Strong collaborative relationships were evident between high-income countries, particularly among Anglo-American and European research partnerships. Lower- and middle-income countries, including India, Pakistan, and various African nations, demonstrated connections primarily with high-income country partners rather than substantial South-South collaboration.

### Regional Topic Focus Analysis

Geographic analysis of topic distribution across seven world regions revealed substantial variation in research priorities and thematic emphasis (**Figure 5**). The heatmap analysis demonstrated marked disparities in regional research focus, with topic prevalence ranging from 7.4% to 85.7% across different region-topic combinations, indicating pronounced geographic specialisation and potential research gaps. More detailed analysis can be found in **Appendix 3**.

**Figure 5:**
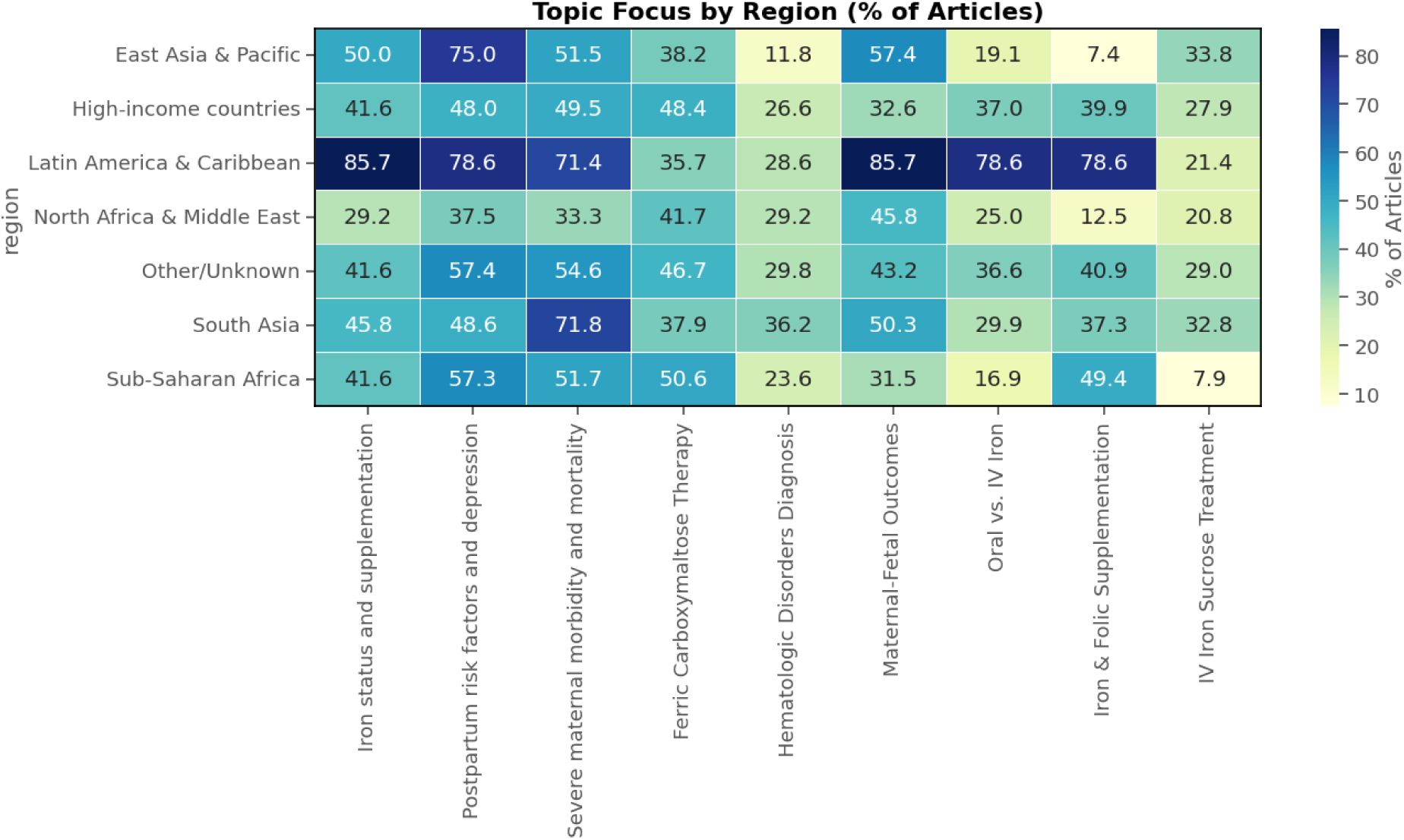
Topic Focus by Region (% of Articles)

### Nine-Cluster Thematic Structure

The topic modelling of the maternal anaemia literature corpus revealed nine distinct thematic clusters (**Table 1 and Appendix 4**). The analysis identified four primary research domains: Prevention (20.8% of documents), Diagnosis (6.3%), Management (21.6%), and Health Outcomes (77.4%), with some documents spanning multiple categories. Comparison with the four categories relevant to guideline development revealed notable distributional differences between computationally-derived clusters and manual classification (**Appendix 5)**. While the clustering identified Health Outcomes as the dominant research focus (77.4%), manual classification showed more balanced distribution across Risk Factors (37.7%), Outcomes (31.2%), and Treatment (28.8%), with Prevention representing only 8.1% compared to the computational clustering’s 20.8%.

**Table 1:** Topic Classification and keywords.

| Cluster | Topic keywords | Research Summary | Number of publications (%) |
| --- | --- | --- | --- |
|  | **<u>Prevention</u>** |  |  |
| #1: Iron status and supplementation | iron status, concentration, status, infant, serum ferritin, vitamin, ferritin, serum, supplement | Nutritional interventions, biomarker assessment, impact of maternal iron deficiency on cognitive health | 150 (14.9) |
| #8: Iron and folic acid supplementation | iron supplementation, supplementation, factor associated, weight gain, acid, severe anaemia, malaria, postpartum period, folic | Anaemia prevention strategies particularly in low-income populations and HIV-infected pregnant women , supplementation efficacy, malaria prophylaxis, policy implications of universal iron supplementation programmes and intervention strategies for improving supplementation coverage in resource-limited settings. | 59 (5.9) |
|  | **<u>Diagnosis</u>** |  |  |
| #5: Thrombocytopenia and postpartum complications | antenatal care, puerperal sepsis, family planning, health facility, management, thalassemia, knowledge, postpartum period, thrombotic thrombocytopenic | awareness gaps regarding anaemia signs in pregnancy, and management, diagnostic awareness of haematological disorders during pregnancy such as atypical haemolytic uraemic syndrome and thrombotic thrombocytopenic purpura | 63 (6.3) |
|  | **<u>Management</u>** |  |  |
| #4: Ferric carboxymaltose and anemia management | trial, ferric carboxymaltose, evidence, quality evidence, blood transfusion, postpartum haemorrhage, primary outcome, quality, blood loss | evidence-based clinical management using intravenous ferric carboxymaltose, blood transfusion reduction, quality evidence assessment, optimal iron delivery methods, comparative effectiveness against standard medical care, and evidence-based guidelines for postpartum anaemia management. | 87 (8.6) |
| #7: Intravenous vs. oral iron therapy | oral iron, intravenous iron, post partum, twin pregnancy, oral, intravenous, twin, blood transfusion, post | clinical impacts of iron deficiency anaemia, therapeutic modalities for postpartum anaemia, comparative effectiveness studies of intravenous and oral iron, standardised treatment protocols, and implementation strategies. | 71 (7.1) |
| #9: Iron sucrose treatment efficacy | third trimester, iron sucrose, trimester, intravenous iron, sucrose, intravenous, oral, oral iron, third | evaluation of intravenous iron sucrose for third-trimester and postpartum anaemia, effectiveness of erythropoietin adjuvant therapy combined with iron sucrose, comparative analyses with oral iron supplementation, and | 59 (5.9) |
| **Health Outcomes** |  |  |  |
| #2: Postpartum risk factors and depression | risk factor, blood loss, postpartum depression, depression, cesarean section, loss, prevalence, cesarean delivery, tranexamic acid | prediction models for postpartum haemorrhage, factors contributing to postpartum depression in various populations, and cell salvage impacts on postpartum anaemia following caesarean delivery. | 250 (24.8) |
| Cluster #3: Severe maternal morbidity and mortality | blood transfusion, maternal death, maternal morbidity, maternal mortality, postpartum haemorrhage, death, caesarean section, obstetric, morbidity | blood transfusion requirements, and caesarean section complications, heart disease-related maternal deaths, severe obstetric morbidity near-miss maternal morbidity in women with anaemia | 326 (32.4) |
| #6: Maternal-fetal outcomes and complications | maternal fetal, birth weight, fetal, hemoglobin level, neonatal, gestational, weight, apgar score, platelet count | thyroid dysfunction on maternal and fetal outcomes in women with anaemia, first-trimester haemoglobin relationships with pregnancy complications. | 203 (20.2) |

### Emerging Research Areas

#### Rapidly Emerging Topics

Two topics demonstrated statistically significant increasing trends with the steepest positive slopes. Ferric carboxymaltose therapy exhibited the most pronounced growth trajectory (slope = +0.156, R² = 0.186, p = 0.027), representing a 134.9% increase between early (2000-2005) and recent (2020-2024) periods. This cluster showed consistent upward progression from approximately 9% prevalence in 2000 to peaks exceeding 15% by 2024. Oral vs. intravenous i ron demonstrated the second-highest growth rate (slope = +0.125, R² = 0.129), reflecting increased comparative effectiveness research between therapeutic modalities.

#### Sustained Growth Areas

Severe maternal morbidity and maternal m ortality maintained steady growth throughout the observation period (slope = +0.085, R² = 0.058, p = 0.246), with prevalence increasing from approximately 12% in 2000 to consistent levels above 15% after 2015. Hematologic Disorders Diagnosis showed moderate positive trends (slope = +0.026, R² = 0.028, p = 0.423), indicating gradual but sustained research expansion in diagnostic approaches.

#### Stable Research Areas

Minimal Change Topics: Several topics such as postpartum depression, risk factors for postpartum anaemia and IV sucrose treatment demonstrated stable and consistent research attention without marked expansion or decline.

### Declining Research Areas

#### Significantly Declining Topics

Iron Status and Supplementation demonstrated the most pronounced decline among all topics (slope = −0.267, R² = 0.287, p = 0.006), showing statistically significant reduction from peaks of approximately 19% in early 2000s to below 10% by 2024. This represented a 24.9% decrease between comparative periods. Maternal-fetal iutcomes showed moderate decline (slope = −0.015, R² = 0.015, p = 0.555), with prevalence decreasing from approximately 15% to below 10% over the study period.

#### Gradual Decline

Iron & folic acid s upplementation exhibited a declining trend (slope = −0.048, R² = 0.048, p = 0.292), demonstrating reduced research focus on traditional supplementation approaches, decreasing from peaks above 15% to approximately 10% prevalence. This decline reflected a 15.3% reduction in research attention over the comparative periods.

## Discussion

Our bibliometric analysis shows that there was exponential publication growth on maternal anaemia increasing nearly ten-fold from 12 publications in 2000 to 118 in 2022. While our bibliometric analysis shows that there has been intensified academic attention to maternal anaemia as a priority area, globally there has been insufficient progress made on reducing anaemia in the last decade.

Understanding the extent and distribution of anaemia and contextualising it to specific settings is crucial to accelerate action on anaemia. The 2021 Global Burden of Disease found that there were large variations in anaemia burden by age, sex, and geography, with women, and countries in sub-Saharan Africa and south Asia being particularly affected. The bibliometric analysis revealed pronounced geographic imbalances contradicting global disease burden patterns. High-income countries dominated research output with 68.5% of publications (United States 41.7%, United Kingdom 26.8%), despite lower anaemia prevalence compared to affected regions. Regional research priorities demonstrated significant specialisation patterns that may not align with local health needs.

To move forward progress on maternal anaemia, the Comprehensive Framework for Action to Accelerate Anaemia Reduction emphasizes a broad approach to diagnosis, prevention and management. Addressing both the causes and risk factors simultaneously is essential for effective anaemia control. However, our bibliometric analysis shows that only about 20% of studies focused on prevention. The studies focused on anaemia prevention strategies in low-income populations, in HIV-infected pregnant women and on malaria prophylaxis. A recent systematic review on the cost effectiveness of interventions for the screening, diagnosis and management of anaemia identified several cost-effective antimalarial regimen and cost-effective delivery channels of antimalarials as well as non-pharmacological interventions. Malaria related anaemia highlights that effective anaemia prevention and management require the development and implementation of contextualized multisectoral strategies, with actions implemented synchronously to address the determinants of anaemia.

There remains a need to clarify the optimal screening method, dosing regimen, timing and route of iron supplementation in pregnancy. The WHO recommendations on antenatal care for a positive pregnancy experience identified priority research questions related to iron including efficacy, acceptability, feasibility, absorption and ideal compound and formulation. However, the bibliometric analysis found that iron status and supplementation demonstrated the most pronounced decline among all topics. Iron and folic acid supplementation also exhibited a declining trend demonstrating reduced research focus on traditional supplementation approaches. On the other hand, Ferric carboxymaltose therapy exhibited the most pronounced growth trajectory. Oral vs. intravenous iron demonstrated the second-highest growth rate.

Our results show that the field has evolved towards treatment-focused and outcome evaluation research with little on prevention, diagnosis and implementation. While sustained attention on prevention and diagnostic research is needed, new evidence on health outcomes can inform recommendations on treatment. Effective anaemia programming requires evidence-based, data-driven, contextualized multisectoral strategies with coordinated implementation. A lack of implementation research hinders the effective scaling of anaemia interventions, especially in low resource settings. Limited implementation research means less is known about barriers to intervention delivery and uptake in real-world settings

As the causes of anaemia are multifactorial and includes micronutrient deficiencies, infections, inflammation, chronic diseases, and inherited blood disorders, the Comprehensive Framework advocates for coordinated action across systems and multisectoral approaches that address both biological and social determinants. To date, there has been limited research on integrated anaemia intervention delivery, despite the complex etiology of anaemia and recommendations for a multisectoral response. Our analysis found that there is a lack of interdisciplinary research. The majority of papers are situated within the field of Obstetrics & Gynaecology. Public health accounted for about 10% of the papers with contributions increasingly markedly after 2010. Other disciplines included General Internal Medicine (14.9%), Haematology (8.9%), and Nutrition & Dietetics (8.4%). While the analysis did not specifically examine collaboration between disciplines, it examined the relationship between topic clusters highlighting opportunities for potential interdisciplinary research.

### Study Strengths and Limitations

This study demonstrates several methodological strengths that enhance the validity and comprehensiveness of findings. The integration of multiple analytical approaches, including traditional bibliometric indicators and advanced topic modelling using Latent Dirichlet Allocation (LDA) provides triangulated evidence strengthening interpretive confidence. This methodological pluralism aligns with best practices in bibliometric research advocated by Donthu et al. (2021)^23^, enabling comprehensive examination of research landscape complexity beyond single-method limitations.

The substantial dataset encompassing 1,007 documents across a 25-year timespan (2000-2024) provides robust temporal perspective for identifying meaningful trends and evolution patterns. This longitudinal scope enables detection of significant changes in research priorities whilst maintaining sufficient sample size for statistical trend analysis. The global coverage spanning 84 countries and 688 institutions offers comprehensive geographic representation, facilitating identification of research concentration patterns and geographic disparities that would remain invisible in smaller or regionally-focused analyses.

The study’s examination of research-burden inversion through geographic analysis addresses critical equity considerations often neglected in bibliometric research. By systematically comparing research output concentration with global disease burden patterns, this analysis provides evidence for research justice arguments and highlights structural inequities in knowledge production systems.

Limitations include the reliance on Web of Science Core Collection as the primary data source introduces potential selection bias, as this database exhibits known coverage limitations for publications from low- and middle-income countries and non-English language journals (Mongeon & Paul-Hus, 2016)^24^. This limitation may have resulted in underrepresentation of research conducted in high-burden regions, potentially exacerbating the observed geographic concentration patterns.

The restriction to English-language publications introduces language bias that may exclude relevant research published in local languages, particularly in regions with high anaemia burden where local language publishing traditions remain strong.

The LDA topic modelling approach, whilst providing objective thematic clustering, inherently reduces complex research content to word co-occurrence patterns, potentially obscuring nuanced conceptual relationships and contextual meaning. As noted by Blei (2012)^25^, topic models represent statistical abstractions that may not fully capture semantic complexity or disciplinary knowledge structures. The manual assignment of topic labels introduces subjective interpretation that, whilst informed by domain expertise, may not reflect universal consensus on thematic boundaries.

The bibliometric approach cannot assess research quality, methodological rigour, or practical impact beyond citation metrics, which exhibit known limitations including citation bias towards certain study types and geographic regions ^26^. This constraint prevents evaluation of whether research concentration in high-income countries produces higher quality or more applicable evidence for addressing maternal anaemia in affected populations.

## Conclusion

While there has been a nearly ten fold exponential growth in publication output on maternal anaemia, this comprehensive bibliometric analysis of maternal anaemia research reveals a dynamic but structurally imbalanced research landscape that fails to align with global health priorities and disease burden distribution. The research landscape shows pronounced geographic concentration of publications in high-income countries despite minimal disease burden, whilst regions most affected by maternal anaemia remain systematically underrepresented in knowledge production. This inversion of research capacity and health need perpetuates knowledge hegemony and limits the development of contextually appropriate and cause specific interventions. The collaborative networks, whilst extensive, reinforce existing power structures rather than fostering South-South knowledge exchange or capacity building in high-burden regions.

Thematically, the field has evolved towards treatment-focused and outcome evaluation research while there is little on prevention, diagnosis and implementation. This provides an opportunity to capitalize on new evidence to develop recommendations on treatment strategies. However, sustained attention is needed on prevention and diagnosis for recommendation generation on these domains but also to support integrated recommendations for treatment and outcomes. Coordination and collaboration is critical for a comprehensive approach to maternal anaemia and our analysis shows that there is limited interdisciplinary connectivity. The lack of multi-disciplinary research translates to a lack of multi-component interventions that are necessary to address maternal anaemia.

## Data Availability

All data produced in the present work are contained in the manuscript

## Annexes

**eFigure 1:**
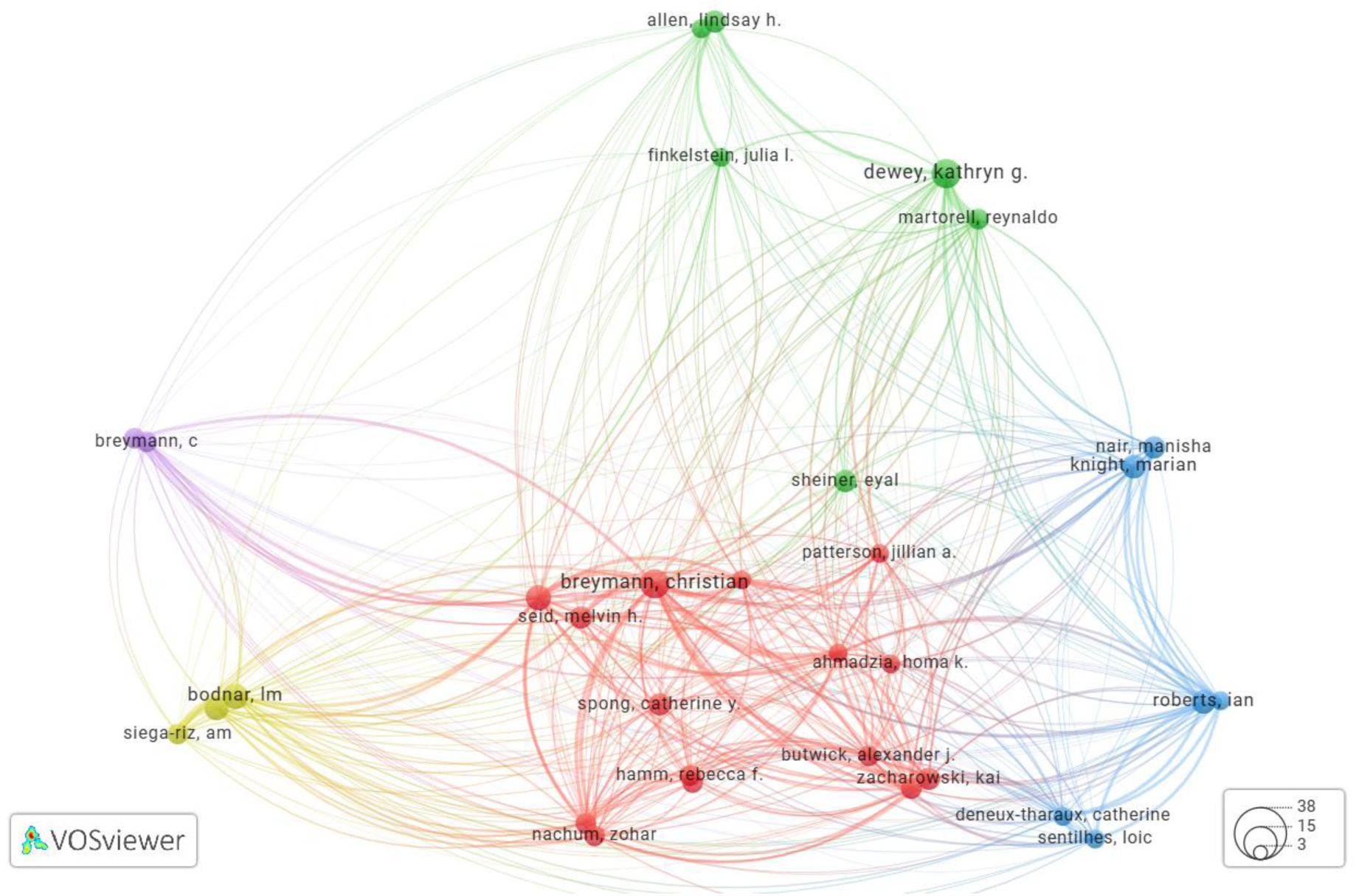
Authors’ collaborations network.

**eFigure 2:**
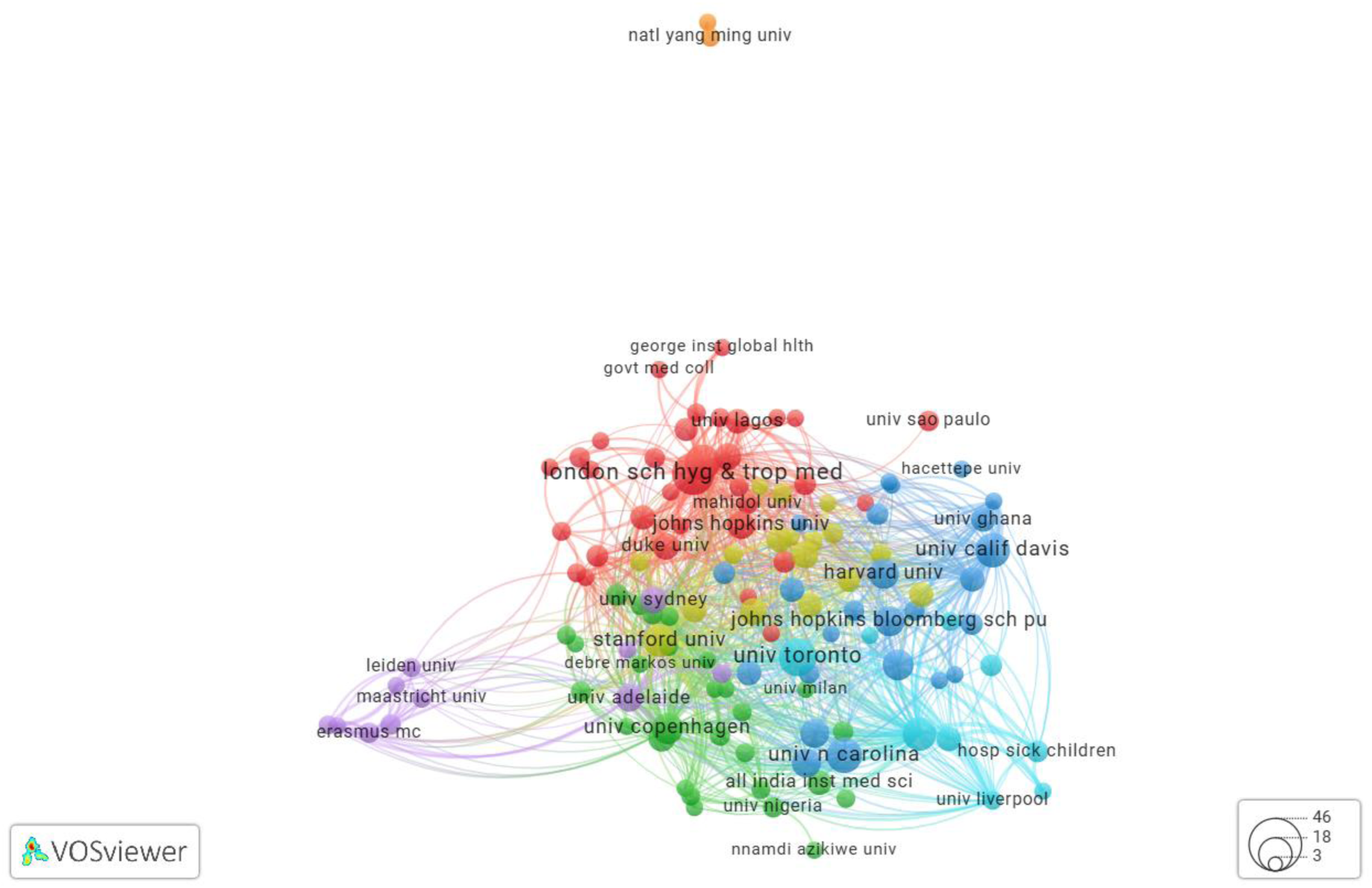
Institutions’ collaborations network.

**eFigure 3:**
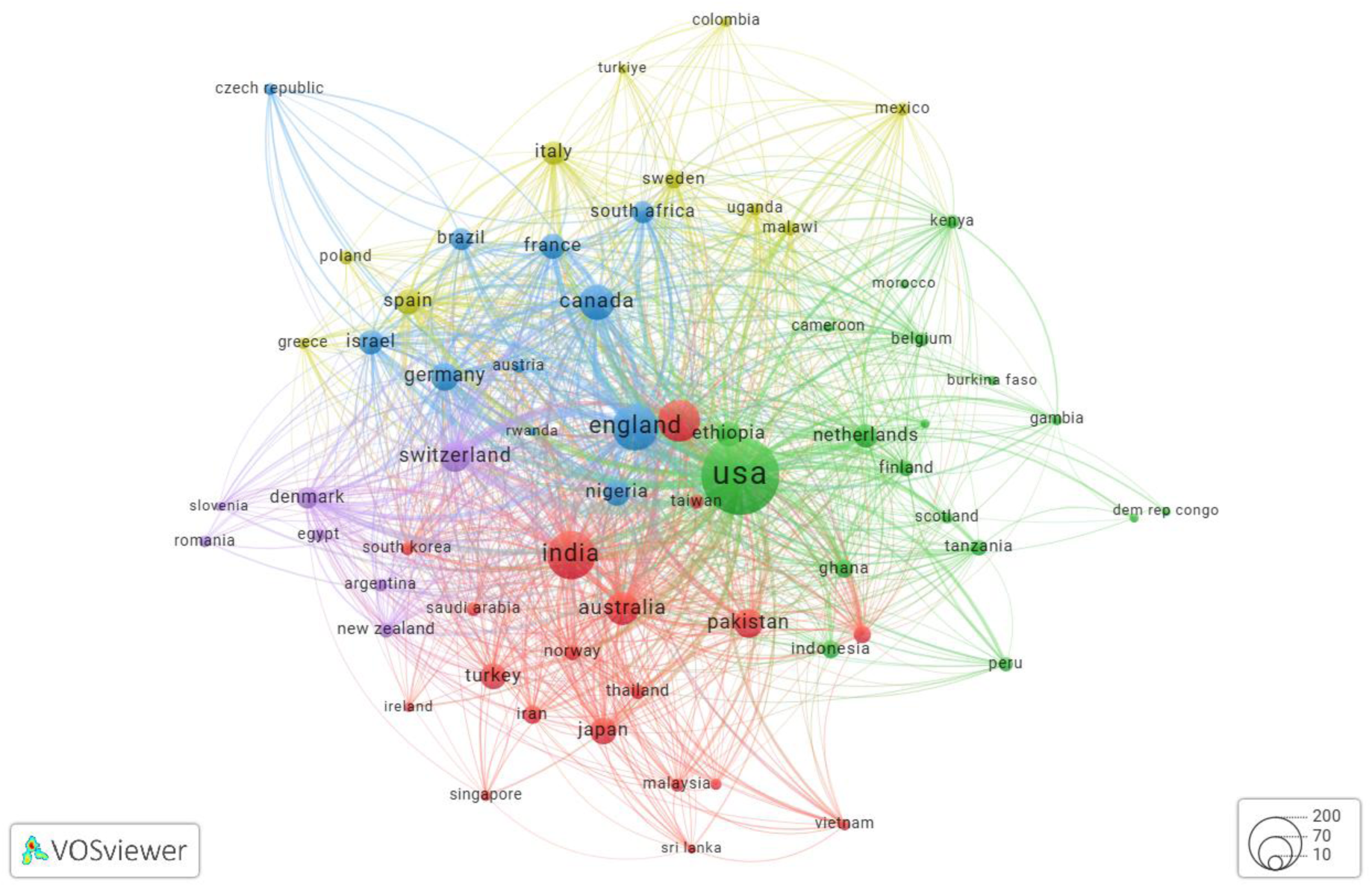
Countries’ collaborations network.

## Appendix 1 METHODS

### Dataset Collection (Step 1)

#### Database Selection and Search Strategy

The Web of Science Core Collection served as the sole bibliographic database for this analysis, selected for its comprehensive coverage of high-impact journals and robust metadata quality essential for advanced bibliometric analysis. A systematic four-phase search strategy was implemented to ensure comprehensive yet focused dataset construction.

Phase 1: Initial Broad Search commenced with an extensive query designed to capture all potentially relevant literature on maternal anaemia. The search strategy employed a combination of Medical Subject Headings (MeSH) terms and free-text keywords, including variations of “anaemia,” “anemia,” “iron deficiency,” combined with pregnancy-related terms such as “pregnant,” “pregnancy,” “postpartum,” “maternal,” and “perinatal.” This initial search yielded 18,768 citations spanning the complete temporal range from 2000 to 2024.

Phase 2: Query Refinement involved systematic narrowing of search parameters through exclusion criteria designed to eliminate studies unrelated to maternal anaemia in pregnancy. Exclusion terms included “chemotherapy,” “oncology,” “malignancy,” “surgery,” and other clinical contexts not relevant to maternal anaemia in pregnancy. This refinement process reduced the dataset to 1,529 citations.

Phase 3: Manual Screening and Selection comprised comprehensive title and abstract screening conducted by reviewers using predefined inclusion and exclusion criteria. Inclusion criteria required studies to focus specifically on anaemia in pregnant or postpartum women, including epidemiological investigations, intervention studies, diagnostic research, and outcome assessments. Exclusion criteria eliminated studies addressing anaemia in non-pregnant populations, paediatric anaemia unrelated to maternal status, and anaemia secondary to non-obstetric causes. This screening process resulted in 1,057 included studies, representing a 69.1% inclusion rate from the refined search results.

Phase 4: WHO Framework Classification involved systematic manual labelling of included studies according to the six WHO Framework categories: Determinants (risk factors and causation), Diagnosis (biomarkers and diagnostic approaches), Prevention (supplementation and preventive interventions), Treatment (therapeutic management), Outcomes (maternal and neonatal health consequences), and Implementation (health systems and policy integration). Multiple reviewers independently classified each study, with disagreements resolved through consensus discussion. Additional 50 studies were excluded at this stage.

Dataset Pre-processing and Refinement (Step 2)

#### Text Corpus Preparation

Comprehensive bibliographic data extraction captured titles, abstracts, author keywords, Keywords Plus, publication years, author affiliations, and citation information for all included studies. Text preprocessing employed natural language processing techniques including stop word removal using an expanded English stop word list supplemented with domain-specific terms (e.g., “study,” “research,” “analysis”). Tokenisation decomposed text into individual terms, whilst lemmatisation reduced words to their root forms to ensure semantic consistency across variant spellings and grammatical forms.

#### Keywords Corpus Generation

A comprehensive vocabulary corpus was constructed from author keywords, Keywords Plus terms, and frequently occurring terms extracted from titles and abstracts. Term frequency analysis identified dominant concepts, whilst rare terms (appearing in fewer than five documents) were excluded to reduce noise. The resulting keywords corpus served as the foundation for semantic network analysis and topic model vocabulary construction.

#### Refinement and Normalisation

Textual data underwent standardisation procedures including acronym expansion, British-American spelling normalisation, and medical terminology standardisation to ensure consistency across the corpus. Author affiliation data were cleaned and standardised to enable accurate institutional and country-level analysis, with institutional name variants consolidated under primary institutional identifiers.

#### Weighting Scheme

Term frequency-inverse document frequency (TF-IDF) weighting was applied to enhance the significance of terms that appear frequently within individual documents but relatively rarely across the entire corpus. This weighting scheme improved the specificity of topic modelling by emphasising distinctive rather than common vocabulary, thereby enhancing the interpretability of extracted topics and reducing the influence of generic academic language.

### Data Analysis (Step 3)

#### Descriptive Analysis

Publication frequency analysis calculated annual publication counts, cumulative growth patterns, and exponential growth modelling using least-squares regression. Geographic distribution assessment examined publication counts by country and region, normalised by population and gross domestic product to identify research intensity patterns. Institutional productivity analysis ranked universities and research centres by publication output whilst controlling for institution size and research capacity. Journal analysis assessed publication venues, impact factors, and disciplinary distribution to characterise the academic landscape surrounding maternal anaemia research.

#### Bibliometric Network Analysis

Co-authorship Network Construction employed VOSviewer software to visualise collaborative relationships among authors, institutions, and countries. Network parameters included minimum publication thresholds (authors: 3 publications; institutions: 5 publications; countries: 10 publications) to ensure network clarity whilst maintaining comprehensive coverage. Centrality measures (degree, betweenness, closeness) identified influential nodes and network bridges facilitating knowledge transfer. Institutional and Country Collaboration Networks mapped international research partnerships and institutional linkages, weighted by collaboration frequency and adjusted for geographic distance to identify preferential collaboration patterns. Network clustering algorithms detected research communities and disciplinary boundaries within the maternal anaemia research landscape.

#### Topic Modelling

Latent Dirichlet Allocation Implementation utilised the gensim Python library to identify latent thematic structures within the text corpus. Document preprocessing included additional medical terminology normalisation and phrase detection to capture multi-word concepts (e.g., “postpartum haemorrhage,” “iron deficiency”). The LDA algorithm assumed each document contained multiple topics in varying proportions, enabling nuanced thematic classification.

Optimal Topic Number Determination employed systematic evaluation of topic coherence scores and model perplexity across topic numbers ranging from 5 to 15. Coherence scores measured the semantic consistency of topic-defining words, whilst perplexity assessed model fit to the data. The optimal number of topics was selected based on coherence score maximisation and human interpretability assessment by domain experts.

Temporal Topic Analysis tracked topic prevalence across the 25-year study period using document-topic probability distributions aggregated by publication year (**Annex 1**). Linear regression analysis identified topics with statistically significant increasing or decreasing trends, whilst change-point detection algorithms identified periods of rapid thematic shifts. Topic evolution visualisation employed stream graphs to illustrate temporal dynamics and research focus transitions.

### Gap Analysis Framework

#### Two-Dimensional Thematic Mapping

The gap analysis framework employed a two-tiered strategy combining computational topic detection with expert-informed WHO Framework classification. Computer-generated LDA topics were systematically mapped onto WHO Framework categories through content analysis of topic-defining keywords and representative documents. This mapping process identified areas of topic overlap, WHO Framework categories lacking corresponding computational topics, and computational topics spanning multiple WHO Framework domains.

### Trend Analyses Methodology Summary

#### A: Methodology

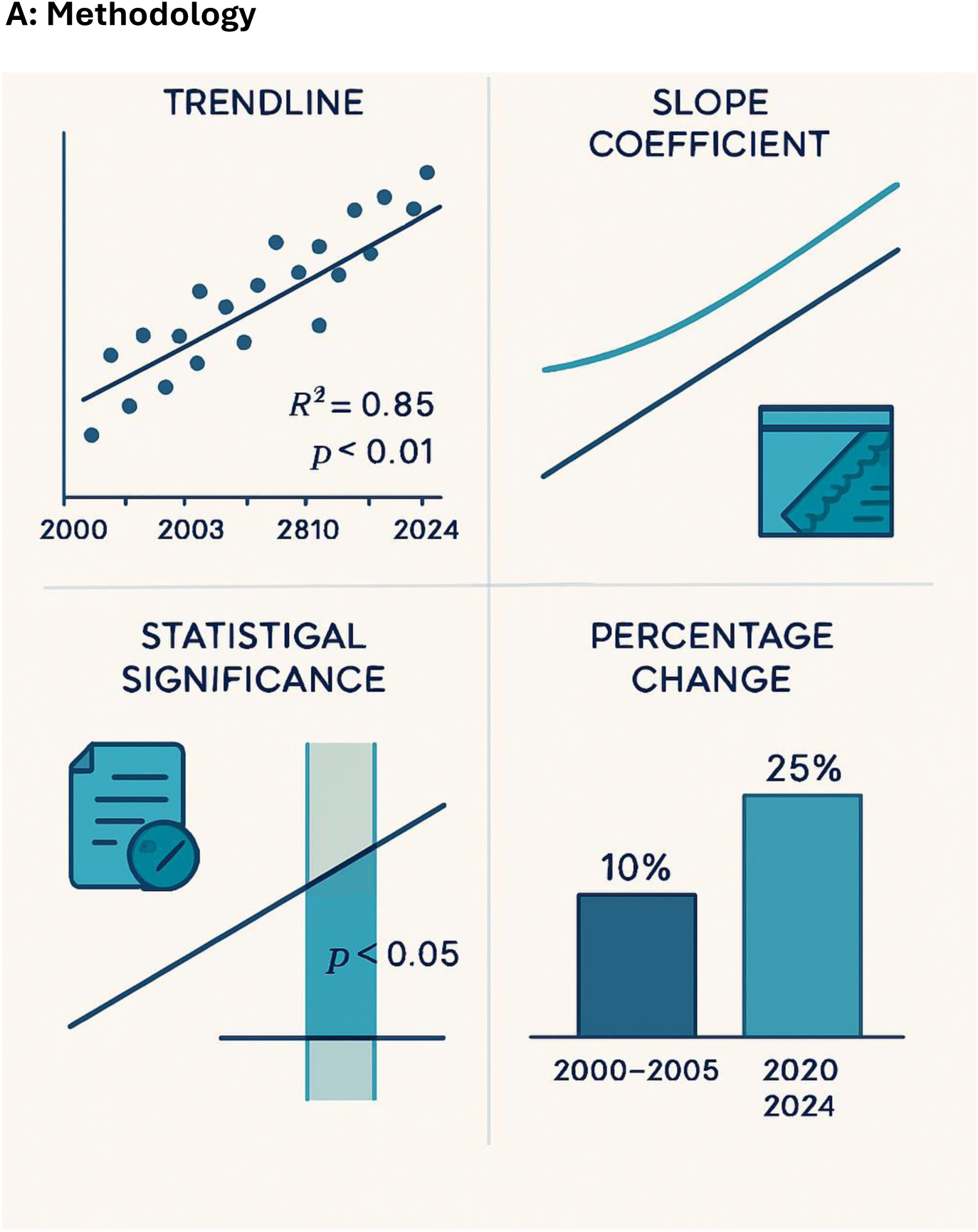
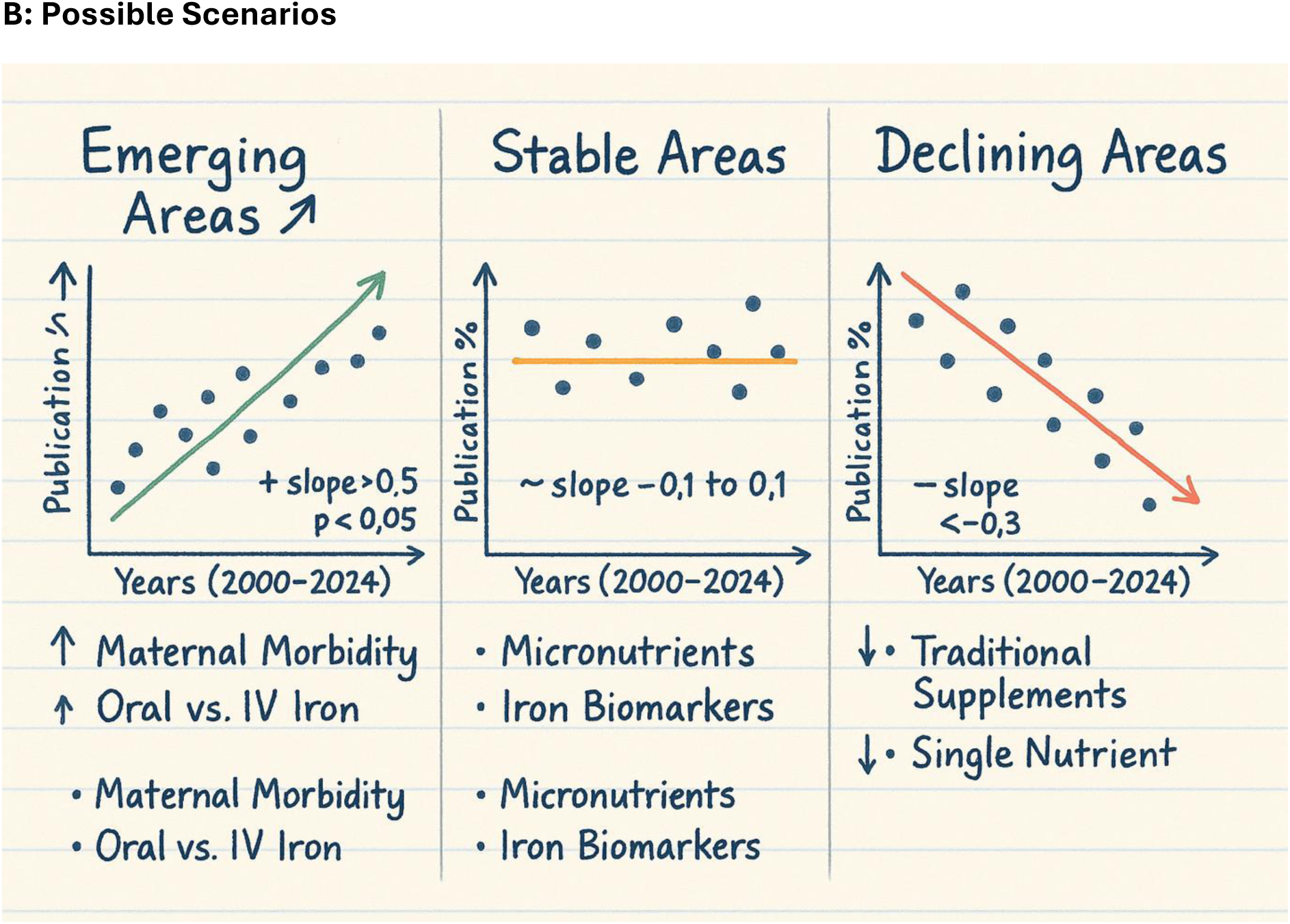

#### B: Possible Scenarios

##### Regional and Temporal Pattern Assessment

Cross-Regional Comparison Analysis calculated topic and WHO Framework category prevalence within seven global regions: Sub-Saharan Africa, North Africa & Middle East, South Asia, East Asia & Pacific, Europe & Central Asia, Latin America & Caribbean, and North America. Regional research intensity was normalised by population and anaemia burden estimates to identify research gaps relative to health need.

Linear Regression Trend Analysis assessed directional changes in research focus through temporal regression analysis of topic and WHO Framework category prevalence. Slope coefficients quantified annual percentage change rates, whilst statistical significance testing (p < 0.05) identified meaningful trends versus random fluctuations. Trend analysis employed robust regression techniques to minimise the influence of outlier years and publication irregularities.

Research Priority Evolution Assessment examined shifts in research emphasis by comparing early period (2000-2005) and recent period (2020-2024) topic distributions. This temporal comparison identified emerging research areas, declining research domains, and stable research focuses, enabling projection of future research trajectory and identification of sustained versus transient research priorities.

##### Interactive Dashboard Development

The dashboard architecture incorporated multiple interconnected modules enabling users to explore different dimensions of the maternal anaemia in pregnancy research landscape. Data preprocessing for dashboard integration involved standardisation of all analytical outputs into compatible formats, including network data serialisation, topic model parameter storage, and temporal trend data structuring. Interactive visualisation libraries including Plotly, NetworkX, and custom JavaScript components were integrated to enable dynamic data exploration with real-time filtering and drill-down capabilities.

##### Data exploration dashboard screenshot

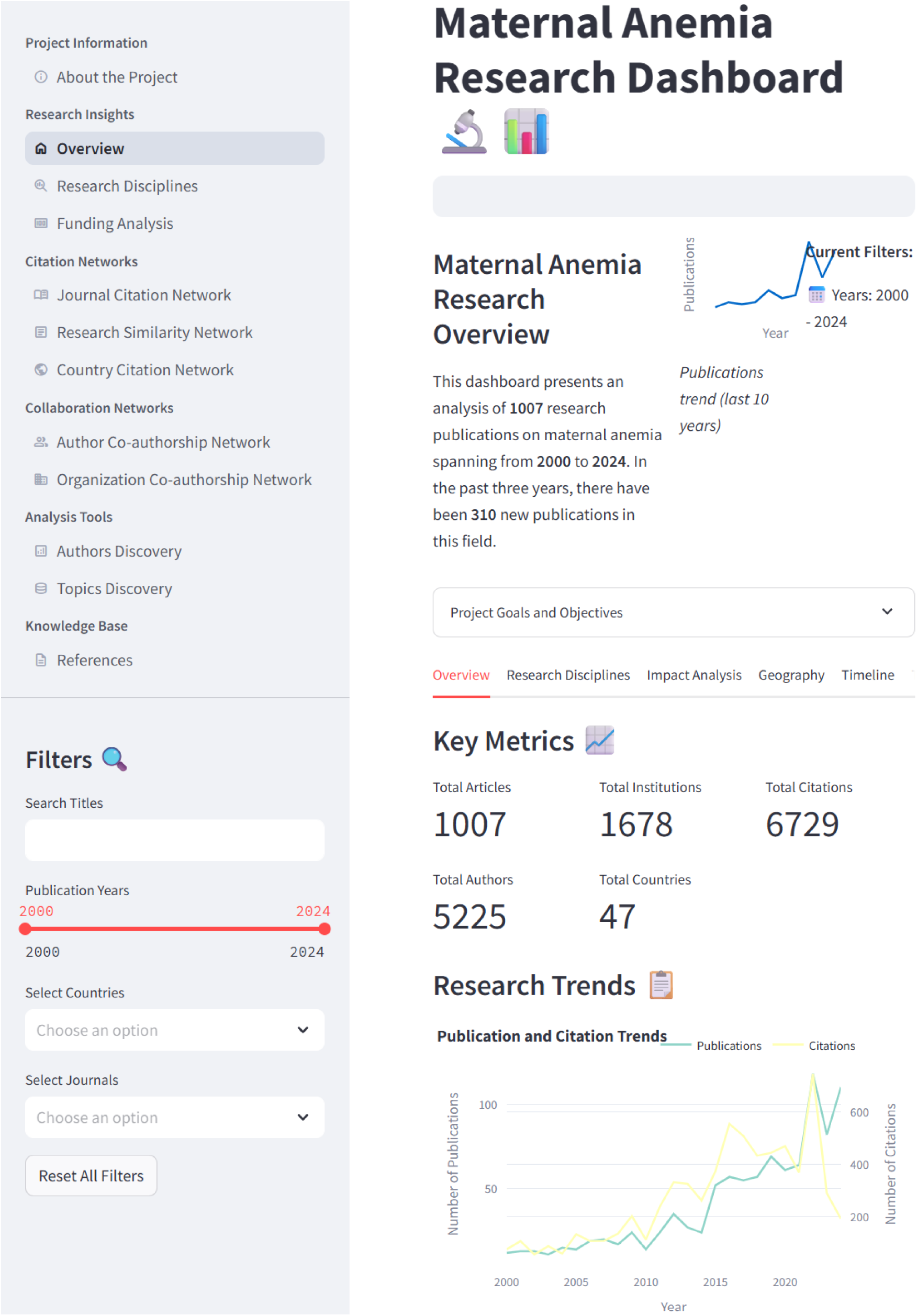

## APPENDIX 2

### Authorship and Collaboration Patterns

The research landscape demonstrated substantial collaborative engagement, with 967 multi-authored documents (96.0%) compared to 40 single-authored publications (4.0%). A total of 5,222 authors contributed to the corpus, yielding an average of 1.16 documents per author and an average collaboration index of 5.82. Author-generated keywords numbered 1,728, whilst Keywords Plus totalled 1,441. Institutional productivity averaged 8.62 documents per institution, and source productivity averaged 2.77 documents per journal or publication venue.

### Author Productivity and Collaboration Networks

Individual author productivity analysis identified ten researchers with the highest publication output in maternal anaemia research (**eFigure 1**). Breymann, Christian emerged as the most prolific author with 12 publications, followed by Bodnar, LM with 10 publications. Dewey, Kathryn G. contributed 9 publications, whilst Milman, Nils and Roberts, Ian each authored 8 publications. Five researchers shared equal productivity with 7 publications each: Cogswell, ME; Sheiner, Eyal; Seid, Melvin H.; Knight, Marian; and Nair, Manisha.

The author collaboration network visualisation revealed distinct research clusters and collaborative partnerships. The network demonstrated a core-periphery structure with Breymann, Christian positioned centrally, indicating extensive collaborative relationships. Notable collaboration clusters included partnerships between Dewey, Kathryn G. and associated researchers, and a separate cluster centred around Nair, Manisha and Knight, Marian. The network displayed moderate density with 38 nodes and multiple interconnected pathways, suggesting active cross-institutional collaboration within the maternal anaemia research community.

### Institutional Distribution and Networks

Institutional productivity analysis revealed concentrated research output among leading academic centres (**eFigure 2**). The University of Toronto demonstrated the highest institutional productivity with 58 publications, followed closely by Harvard University with 52 publications. The University of London contributed 40 publications, whilst Université Paris Cité and the University of North Carolina Chapel Hill each produced 39 publications. The University of California System generated 38 publications, and the Harvard T.H. Chan School of Public Health and Assistance Publique Hôpitaux Paris (APHP) each contributed 33 publications. The London School of Hygiene & Tropical Medicine and the University of North Carolina completed the top ten with 32 publications each.

The institutional collaboration network demonstrated extensive international partnerships and research connectivity. The network revealed three primary institutional clusters: a North American cluster centred around Harvard University, University of Toronto, and Johns Hopkins University; a European cluster featuring the London School of Hygiene & Tropical Medicine and associated UK institutions; and an Asia-Pacific cluster including institutions from India, Australia, and other regional partners. The network structure indicated 46 nodes with substantial interconnectivity, reflecting robust international collaboration in maternal anaemia research.

## APPENDIX 3 Geographic Distribution and Regional Research Patterns

### High-Focus Regional Specialisations

Latin America & Caribbean demonstrated the most pronounced regional specialisation patterns, exhibiting exceptionally high focus on specific topics. Iron Status and Supplementation dominated regional research with 85.7% prevalence, representing the highest topic concentration observed across all regions. Maternal-Fetal Outcomes similarly achieved 85.7% regional prevalence, whilst Iron & Folic Supplementation and Oral vs. IV Iron both maintained substantial regional focus at 78.6% prevalence each. This pattern indicated strong regional emphasis on preventive and comparative therapeutic research approaches.

East Asia & Pacific showed concentrated research attention on postpartum complications, with Postpartum Risk Factors and Depression achieving 75.0% regional prevalence. Maternal-Fetal Outcomes reached 57.4% prevalence, whilst Severe Maternal Morbidity and Mortality maintained 51.5% focus. The region demonstrated balanced attention across outcome-focused research domains.

South Asia exhibited concentrated focus on severe maternal complications, with Severe Maternal Morbidity and Mortality achieving 71.8% regional prevalence. Maternal-Fetal Outcomes maintained 50.3% prevalence, whilst Postpartum Risk Factors and Depression reached 48.6% regional focus, indicating priority attention to acute maternal health outcomes.

### Moderate Regional Concentrations

High-Income Countries demonstrated relatively balanced topic distribution with moderate concentrations across multiple domains. Severe Maternal Morbidity and Mortality achieved 49.5% prevalence, whilst Postpartum Risk Factors and Depression and Ferric Carboxymaltose Therapy both maintained 48.0% and 48.4% prevalence respectively. This pattern suggested diversified research portfolios with broad thematic coverage.

Sub-Saharan Africa showed mixed research priorities with Severe Maternal Morbidity and Mortality achieving 51.7% prevalence and Postpartum Risk Factors and Depression reaching 57.3%. However, the region demonstrated substantial variation across other topics, with some showing moderate focus and others exhibiting significant gaps.

### Pronounced Regional Research Gaps

Limited Research Focus Areas: Several regions demonstrated markedly low research attention in specific domains, indicating potential research gaps. Sub-Saharan Africa exhibited minimal focus on IV Iron Sucrose Treatment (7.9%) and Oral vs. IV Iron (16.9%), suggesting limited attention to advanced therapeutic comparisons despite high anaemia burden. East Asia & Pacific showed restricted focus on IV Iron Sucrose Treatment (7.4%) and Hematologic Disorders Diagnosis (11.8%).

North Africa & Middle East demonstrated the most pronounced research gaps across multiple domains, with IV Iron Sucrose Treatment achieving only 12.5% prevalence and Oral vs. IV Iron reaching 25.0%. Hematologic Disorders Diagnosis maintained 29.2% prevalence, whilst Iron Status and Supplementation achieved only 29.2% regional focus, indicating substantial research underrepresentation across key domains.

Latin America & Caribbean exhibited selective research gaps despite high overall activity, with IV Iron Sucrose Treatment achieving only 21.4% prevalence and Hematologic Disorders Diagnosis reaching 28.6%, suggesting limited attention to specific diagnostic and advanced therapeutic domains.

### Regional Research Intensity Patterns

High-Intensity Regions: Latin America & Caribbean demonstrated the highest research intensity with multiple topics exceeding 70% prevalence, followed by East Asia & Pacific with several topics above 50% prevalence. These regions showed concentrated research efforts with clear thematic priorities.

Moderate-Intensity Regions: High-Income Countries and South Asia exhibited moderate research intensity with balanced topic distribution, suggesting diversified research approaches without extreme specialisation patterns.

Low-Intensity Regions: North Africa & Middle East and Other/Unknown regions demonstrated lower overall research intensity with few topics exceeding 45% prevalence, indicating potential underrepresentation in maternal anaemia research or limited research capacity in specific thematic domains.

The geographic analysis revealed significant research inequities, with some regions demonstrating concentrated expertise in specific domains whilst exhibiting substantial gaps in others, highlighting opportunities for enhanced international collaboration and capacity building in underrepresented areas.

## APPENDIX 4 Nine-Cluster Thematic Structure

### Cluster Analysis Summary

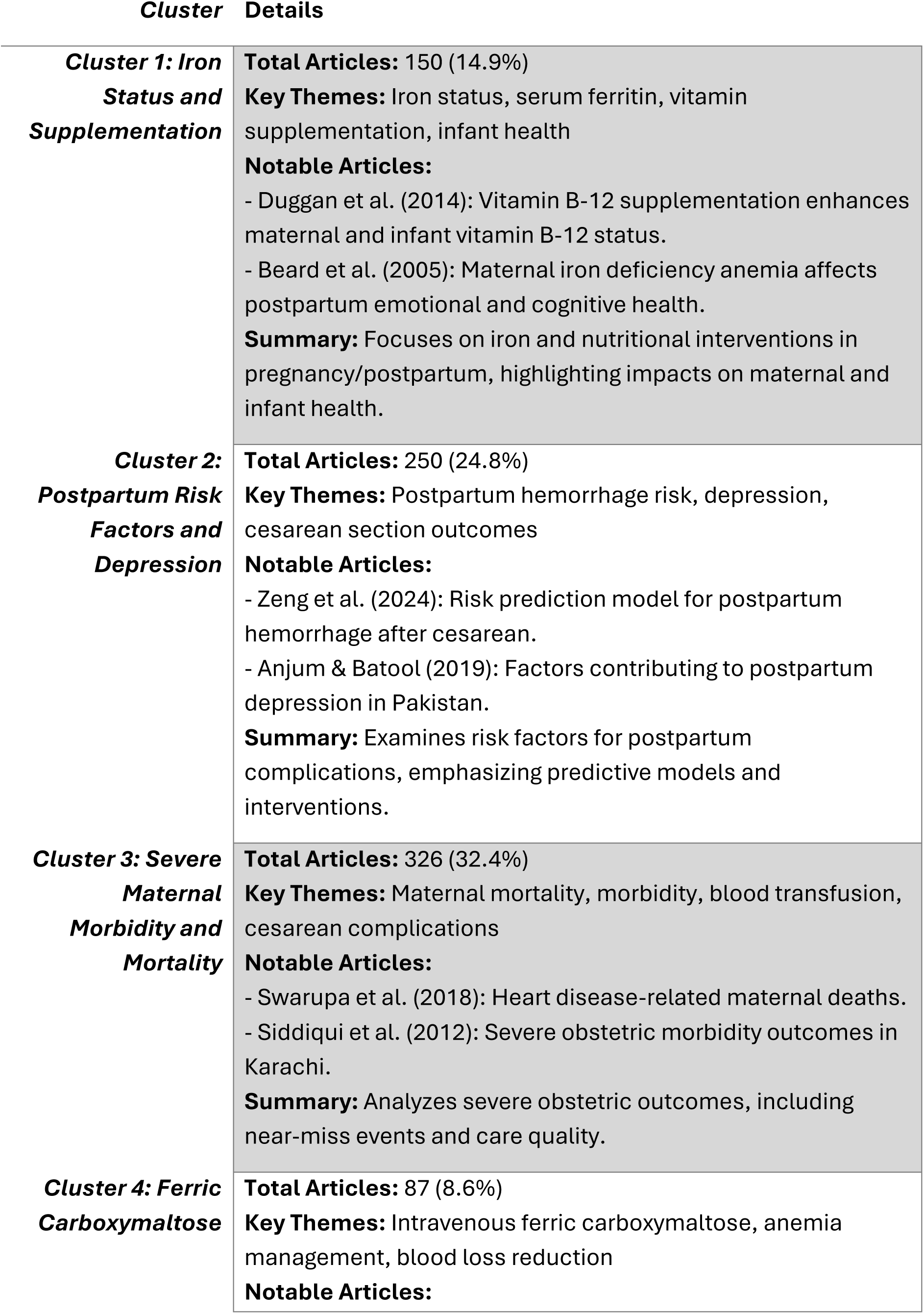
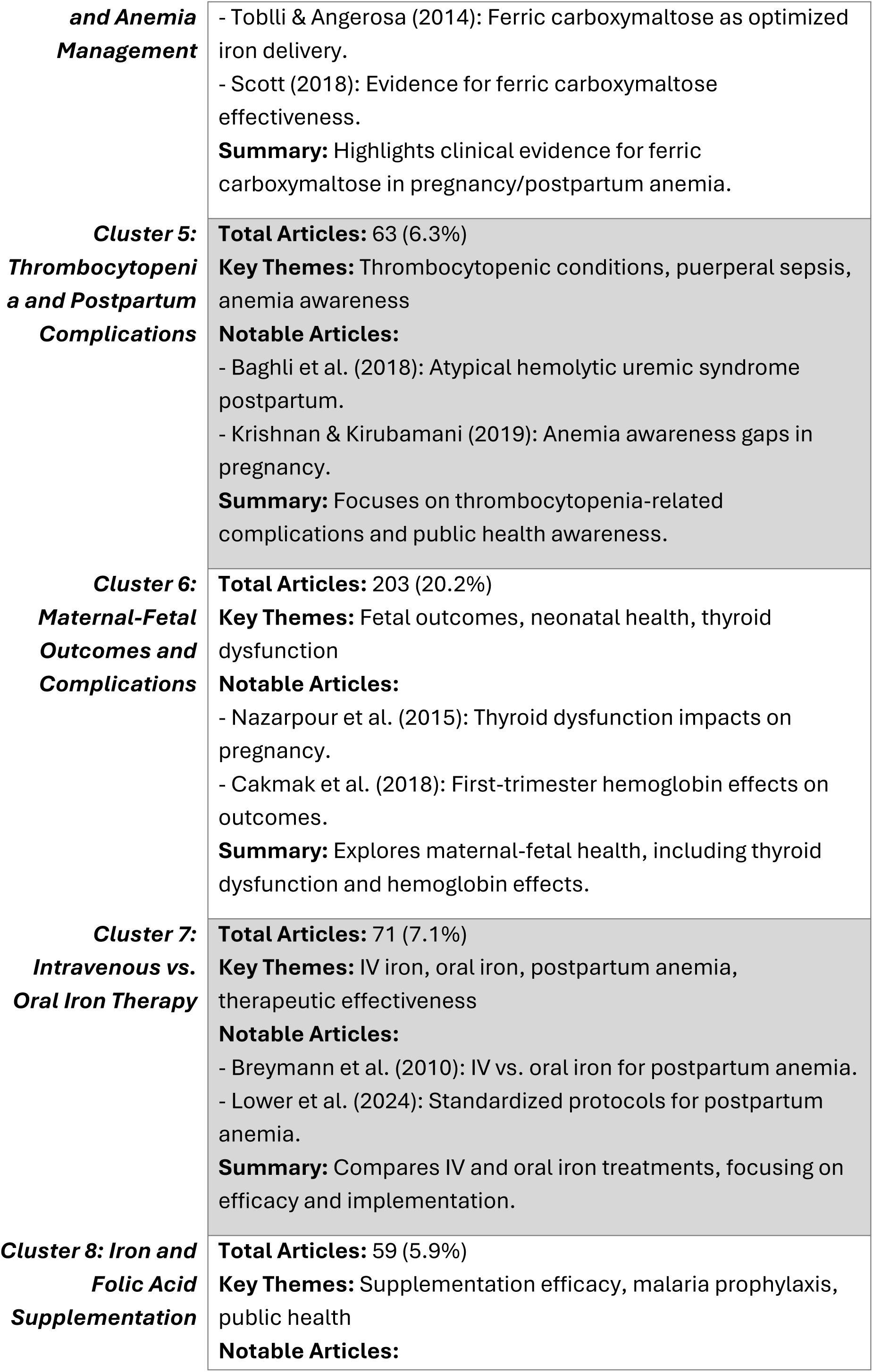
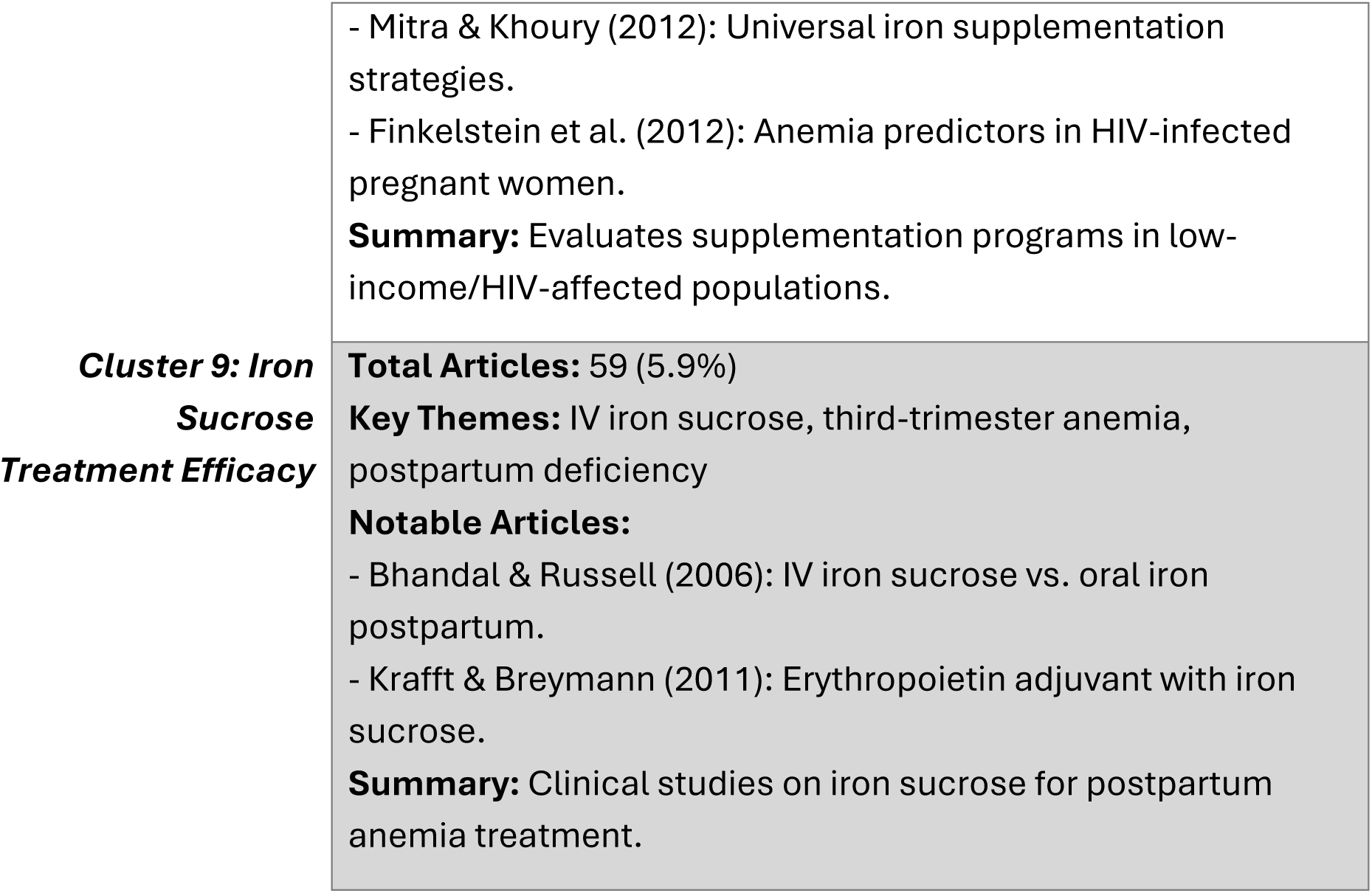

### Network Structure and Co-occurrence Patterns

The topic relationship network revealed complex interdisciplinary relationships among the nine research domains identified through LDA topic modelling, with varying degrees of connectivity and collaborative potential. Network analysis identified three distinct categories based on connectivity patterns: Central Topics (red nodes), Bridging Topics (orange nodes), and Specialised Topics (blue nodes). Node sizes corresponded to topic frequency within the corpus, whilst edge thickness represented co-occurrence strength between topics, with numerical values indicating the number of studies exhibiting dual topic presence.

**Figure 5a:**
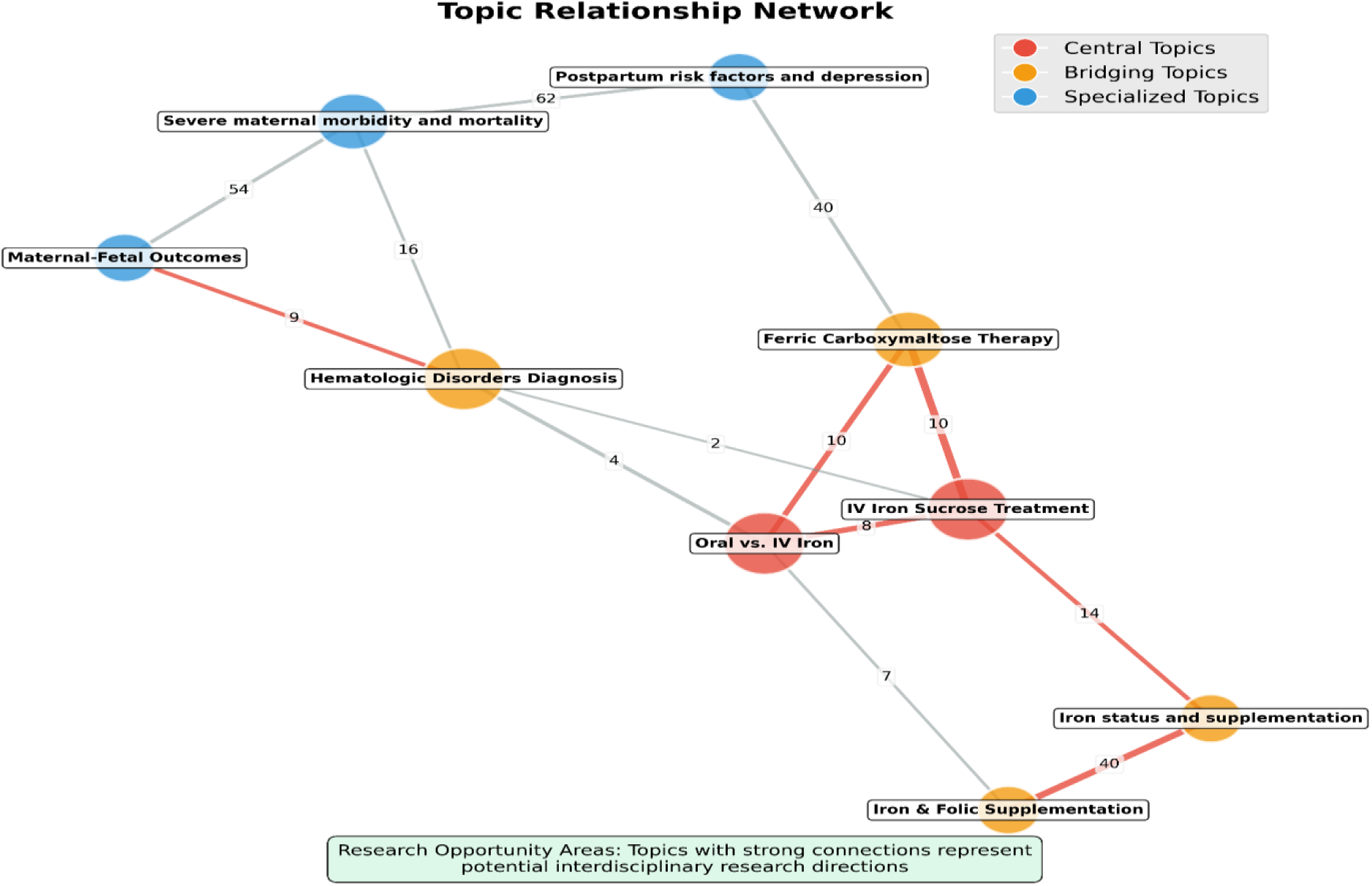
Topic Relationship Network.

Central Topics: IV Iron Sucrose Treatment emerged as the primary central node with the highest connectivity across multiple research domains (64 studies total frequency). This topic demonstrated substantial co-occurrence with Ferric Carboxymaltose Therapy (10 studies), Oral vs. IV Iron (10 studies), and Iron Status and Supplementation (14 studies), indicating its fundamental role in connecting diverse therapeutic approaches within maternal anaemia literature. The central positioning reflected IV Iron Sucrose Treatment’s integration with both comparative effectiveness research and broader supplementation strategies. Oral vs. IV Iron functioned as another central topic (53 studies), serving as a critical junction connecting treatment modalities with prevention approaches. This topic demonstrated moderate co-occurrence with Hematologic Disorders Diagnosis (4 studies) and Iron & Folic Supplementation (7 studies), suggesting its role in bridging therapeutic decision-making with diagnostic considerations and preventive interventions.

Bridging Topics: Ferric Carboxymaltose Therapy served as the primary bridging topic (59 studies), facilitating connections between advanced therapeutic approaches and broader research domains. Beyond its strong connection with IV Iron Sucrose Treatment (10 studies), this topic demonstrated significant co-occurrence with Postpartum Risk Factors and Depression (40 studies), indicating integration of advanced iron therapy research with maternal mental health and complication assessment. Hematologic Disorders Diagnosis functioned as a crucial bridging topic (31 studies), connecting diagnostic research with multiple clinical domains. This topic demonstrated connections with Maternal-Fetal Outcomes (9 studies), Severe Maternal Morbidity and Mortality (16 studies), and moderate linkage with Oral vs. IV Iron (4 studies), suggesting its role in linking diagnostic approaches with outcome assessment and treatment selection. Iron Status and Supplementation represented another significant bridging topic (128 studies), connecting prevention-focused research with treatment approaches. This topic demonstrated strong co-occurrence with IV Iron Sucrose Treatment (14 studies) and substantial connection with Iron & Folic Supplementation (40 studies), indicating its role in linking biomarker assessment with therapeutic interventions.

Specialised Topics: Severe Maternal Morbidity and Mortality exhibited selective connectivity patterns (217 studies), maintaining substantial connections with Postpartum Risk Factors and Depression (62 studies) and moderate linkage with Hematologic Disorders Diagnosis (16 studies). This pattern indicated the topic’s specialised focus on acute maternal complications whilst maintaining selective integration with risk assessment and diagnostic research.

Maternal-Fetal Outcomes demonstrated limited connectivity (176 studies) with primary connection to Hematologic Disorders Diagnosis (9 studies) and weak linkage to Severe Maternal Morbidity and Mortality (54 studies). This specialisation suggested focused research attention on pregnancy outcomes with minimal integration across other research domains. Postpartum Risk Factors and Depression showed concentrated connectivity (227 studies) primarily linking with Severe Maternal Morbidity and Mortality (62 studies) and Ferric Carboxymaltose Therapy (40 studies), indicating specialised focus on postpartum complications with selective therapeutic integration.

### Co-occurrence Strength Analysis

High-Strength Connections: The Postpartum Risk Factors and Depression relationship with Severe Maternal Morbidity and Mortality demonstrated the strongest co-occurrence (62 studies), indicating substantial research integration between postpartum complications and severe maternal outcomes. The connection between Severe Maternal Morbidity and Mortality and Maternal-Fetal Outcomes (54 studies) represented another high-strength association, suggesting integrated approaches to maternal and fetal health assessment.

Moderate-Strength Connections: The Iron Status and Supplementation relationship with Iron & Folic Supplementation (40 studies) and Ferric Carboxymaltose Therapy connection with Postpartum Risk Factors and Depression (40 studies) demonstrated moderate co-occurrence, indicating balanced research attention spanning prevention, treatment, and outcome domains.

Emerging Therapeutic Clusters: The treatment-focused cluster comprising IV Iron Sucrose Treatment, Ferric Carboxymaltose Therapy, and Oral vs. IV Iron showed interconnected relationships (10 studies each between primary nodes), suggesting emergence of comparative effectiveness research as a distinct domain within maternal anaemia literature.

### Temporal Trend Analysis of Topic Categories

Linear regression analysis of topic prevalence over the 25-year study period identified significant temporal trends across the nine thematic clusters (**Figure 6**). Trend analysis revealed varying patterns of research evolution, with slope coefficients ranging from +0.156 to −0.267, indicating substantial shifts in research focus and priorities within maternal anaemia literature.

**Figure 6:**
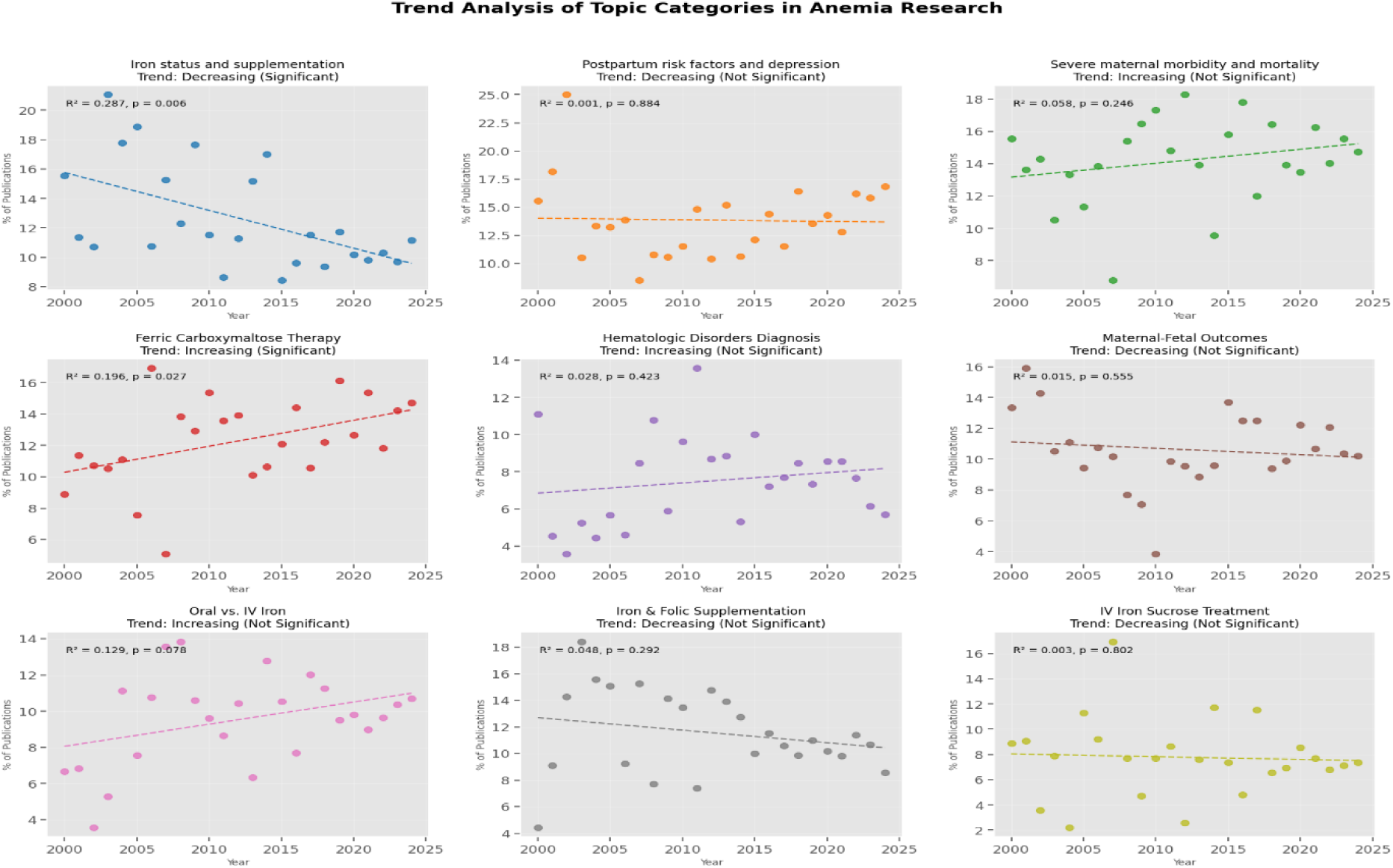
Trend Analysis of Topic Categories.

## APPENDIX 5 WHO Framework Classification

| s/n | Category | Number | Percentage |
| --- | --- | --- | --- |
| **1** | **Risk factors** | **380** | **3.74** |
| 1a | Biological mechanisms (blood loss, haemolysis, ineffective erythropoiesis) | 69 | 6.85 |
| 1b | Direct causes (micronutrient deficiencies; infectious and chronic diseases; gynecological and obstetric conditions, eg PPH; inherited red blood cell disorders) | 264 | 26.22 |
| 1c | Intermediate risk factors (food insecurity, poor diet, lack of access to and use of family planning, limited access to health and nutrition services, inadequate access to WaSH) | 75 | 7.45 |
| 1d | Underlying risk factors (low education, poverty, conflict, cultural norm and behaviors, health policies, socio-demographics, politics, economy, inequity) | 131 | 13.01 |
| **2** | **Diagnosis** | **153** | **15.19** |
| 2a | Biomarkers, lab assessment & QA | 102 | 10.13 |
| 2b | Physical signs & symptoms | 10 | 0.99 |
| 2c | Screening approaches & protocols | 22 | 2.18 |
| 2d | Cause-specific diagnosis (incl. ferritin) | 33 | 3.28 |
| 2e | Point-of-care & innovative diagnostics | 4 | 0.4 |
| 2f | Diagnostic algorithms & decision support | 23 | 2.28 |
| **3** | **Prevention** | **82** | **8.14** |
| 3a | Nutrition (diet, fortification, supplementation) | 64 | 6.36 |
| 3b | Infectious diseases (bednets, deworming, IPTp, WaSH) | 7 | 0.7 |
| 3c | Family planning | 2 | 0.2 |
| 3d | Multi-component interventions | 2 | 0.2 |
| 3e | Education & counselling | 3 | 0.3 |
| 3f | Intervention adherence & compliance | 5 | 0.5 |
| 3g | Management of side effects | 2 | 0.2 |
| **4** | **Treatment** | **290** | **28.80** |
| 4a | Oral micronutrient supplementation | 115 | 11.42 |
| 4b | Intravenous iron | 129 | 12.81 |
| 4c | Blood transfusion | 87 | 8.64 |
| 4d | Treatment of infections (malaria, etc) | 1 | 0.1 |
| 4e | Multi-component interventions | 51 | 5.06 |
| 4f | Treatment adherence & compliance | 6 | 0.6 |
| 4g | Management of side effects | 9 | 0.89 |
| 4h | Treatment monitoring & response | 18 | 1.79 |
| **5** | **Outcomes** | **314** | **31.18** |
| 5a | Impact assessment | 314 | 31.18 |
| **6** | **Implementation** | **14** | **1.39** |
| 6a | Integration in health care systems | 4 | 0.4 |
| 6b | Implementation strategies | 3 | 0.3 |
| 6c | Access & coverage of interventions/services | 9 | 0.89 |

